# The effect of a digital health physical activity program integrating gamification for obesity management in comparison to the usual care: a randomized controlled trial with ideographic approach

**DOI:** 10.1101/2025.02.26.25322872

**Authors:** Alexandre Mazéas, Aïna Chalabaev, Marine Blond, Charline Mourgues, Bruno Pereira, Martine Duclos

**Author notes:** **Author for correspondence:** Alexandre Mazéas. The DOI is available at the top of this page. **Cite this article:** Mazéas, A., Chalabaev, A., Blond, M., Mourgues, C., Pereira, B., Duclos, M. (2025). The effect of a digital health physical activity program integrating gamification for obesity management in comparison to the usual care: a randomized controlled trial with ideographic approach. MedRxiv.^*^. **Data, code, & materials:** DOI 10.17605/OSF.IO/YPD5S.

## Abstract

Digital health interventions and gamification hold promise for managing chronic diseases, but evidence comparing their efficacy to usual care is limited. This two-arm parallel randomized controlled trial with embedded N-of-1 analyses assessed whether a digital intervention integrating gamification and telecoaching (Kiplin) outperform a supervised face-to-face adapted physical activity (PA) program (usual care) in obesity management. Patients with obesity and/or T2D (N = 50, Mage = 46.7 years, 74% female) were randomized to either the Kiplin digital intervention or usual care group, and completed a 3-month program with a 6-month follow-up. The primary outcome was the change in daily step count between baseline and the intervention’s end. Secondary outcomes assessed changes in daily steps, PA, quality of life, body composition, and physical capacities over nine months. Mixed-effects models and generative additive models were conducted to analyze both between- and within-person evolutions in PA. A cost-utility analysis was computed to compare the cost-effectiveness of the programs. Compared to usual care, Kiplin participants showed significant increases in daily steps during the intervention and follow-up periods, with sustained effect sizes. Idiographic analyses revealed variable individual responses, highlighting the need for tailored approaches. No significant differences were observed in secondary clinical outcomes or the cost-utility analysis. This study demonstrates the potential of digital interventions to sustain PA improvements that could offer an alternative to face-to-face programs, but the clinical and economic impacts need to be further evaluated.

## INTRODUCTION

Adapted Physical Activity (APA) is an evidence-based, multidisciplinary rehabilitation program currently considered the gold-standard of treatment of obesity or Type 2 Diabetes (T2D). These programs include physical and sports activities, tailored to the person’s abilities and health condition. APA is considered a true “pillar” in the management of overweight and obesity because of its multiple benefits such as weight and fat loss, improvements in blood pressure, cardiorespiratory fitness, insulin sensitivity, appetite control, and quality of life.^1^ Moreover, the economic impact of such programs could be substantial as, in France, the estimated healthcare savings per individual who becomes sustainably active amount to €840 for individuals aged 20 to 39 and €23,275 for individuals aged 40 to 74.^2^

Despite these well-documented benefits, APA programs often struggle with low adherence and observance rates, with high drop-out rates being reported.^3–5^ Moreover, only a low percentage of patients maintain their physical activity at the end of these programs – participants often failing to maintain regular PA in the long haul.^6–8^ Additionally, logistical issues such as scheduling, travel, childcare, and cost further limit the feasibility and effectiveness of these supervised programs.^9–11^

Given these challenges, there is increasing interest in leveraging digital health technologies for obesity treatment. Digital health interventions hold promise to better change behavior in real-life contexts and have the potential to extend the reach of evidence-based behavioral interventions at lower cost while reducing patient burden.^12^ The interventional perspectives are multiple with the democratization of digital tools. Among these innovations, gamification stands out, defined as the use of game mechanisms such as points, levels, leaderboards, badges, challenges, or customization in non-game contexts to foster behavior change, engagement, motivation, and soliciting participation. Recent meta-analyses suggest that gamified interventions are effective in promoting long-term changes in PA, among various participants, including clinical populations.^13–15^ In addition, telehealth solutions are emerging as a safe alternative for proposing remote APA sessions with good acceptability among patients and positive clinical results, even comparable to conventional face-to-face approaches.^16,17^

Nevertheless, despite the promise of digital health interventions, the evidence supporting their use in obesity treatment remains limited. Although digital interventions have been extensively tested in comparison to passive or active control groups, no rigorous trial has yet demonstrated the superiority of digital PA interventions over existing ones (e.g., supervised and APA programs). As an example, a previous systematic review^18^ revealed that eHealth interventions were effective in promoting PA in adults aged >55 years but did not conclusively demonstrate that eHealth interventions had a greater impact than traditional non-eHealth approaches. Also, evidence regarding long-term effects and cost-effectiveness remains inconclusive.^19,20^ In this context, the digital health domain needs more rigorous evaluations, including follow-up assessments, comparisons with usual care, and economic evaluations.

In addition, although the gold-standard evaluation method, namely the randomized controlled trial (RCT), can be helpful to evaluate the efficacy and effectiveness of digital health tools ^21,22^, evaluation processes must extend beyond the average response in order to consider the idiosyncratic nature of physical activity (i.e., the fact that people’s activity and responses to an intervention can largely differ from one person to another).^23^ This emphasizes the need to adopt an idiographic approach (i.e., individual statistical modeling) and report both between-group differences in benefit and within-person evolutions.^24^ This idiographic approach may help to better explain the heterogeneity frequently observed in the effect of behavioral intervention between different individuals.^25,26^

Finally, as PA behavior change process is dynamic, it requires high-resolution measurements in order to capture its relative variability or stability over time.^23,27^ Since intervention effects can be non-linear and vary across timescales, the traditional measurement bursts conducted in physical activity trials (i.e., 7-day measurement before and after the intervention) are not adapted to properly assess the impact of a behavioral change intervention on physical activity.^23^ To answer these two issues, integrating intensive longitudinal measures with N-of-1 methods within a RCT may prove particularly valuable.^24^

Herein, we present the results of the Digital Intervention Promoting Physical Activity among Obese people (DIPPAO) study, a RCT designed to compare a digital intervention integrating gamification and telecoaching with the usual care. The study tests the hypothesis that these two interventional features can augment PA levels during and at the end of the program – ultimately leading to better clinical outcomes in the long haul. To address limitations observed in previous studies, this trial included: a) a six-month follow-up to assess the sustainability of the intervention effect after the end of the intervention; b) a cost-utility analysis to assess the economic impact of both programs; c) an intensive longitudinal assessment of daily steps throughout the entire study to capture daily PA behavior more precisely; and d) an analysis of both between-group differences and within-person evolutions using an idiographic approach.

## METHODS

### Overview

The DIPPAO study is a two-arm parallel prospective RCT conducted in France (ClinicalTrials.gov NCT04887077, registration date 05/14/2021). Full details of the study methods have been reported previously.^28^ This trial aimed to compare the effectiveness, cost-effectiveness, and psychological mechanisms of the Kiplin program – a group-based digital intervention integrating gamification and supervised PA by telecoaching – in comparison to the usual care (i.e., face-to-face supervised PA sessions) among patients treated for obesity. The authors are solely responsible for the design and conduct of this study, all analyses, drafting, and editing of the paper, and its final contents. The protocol of this trial has been reviewed and approved by the National Human Protection Committee (CPP Ile de France XI, N° 21 004-65219). In this paper, we report the primary and secondary clinical outcomes along with the cost-utility analysis; the psychological mechanisms results are reported separately. There were no significant deviations from the prespecified protocol during the trial. The elements that may differ between the protocol and the final analysis will be specified in the following methods section.

### Study population and procedure

Patients were screened between June 2021 and October 2022. Eligible patients were 18–65 years old who were treated for obesity (BMI ≥30kg/m2 and <45kg/m2) and/or overweight/ obesity and T2D within the department of sports medicine at the University Hospital of Clermont-Ferrand, France. Patients were required to own either an Android-based phone or Apple iPhone with a study supported operating system. Full inclusion and exclusion criteria are available in Table S2 in supplementary materials.

Participants attended five visits throughout the study: a selection visit, an inclusion visit, and three experimental visits. During the selection visit, one of the investigating physicians checked the patients’ ability to complete the full protocol based on eligibility criteria and signed the informed consent form. Randomization was performed by the associate biostatistician using a permuted block design with variable block sizes and a 1:1 allocation ratio. The T0 experimental visit marked the baseline assessment conducted prior to the start of the intervention. At the conclusion of the 3-month program, participants completed the T1 experimental visit. To evaluate the long-term effects of the intervention, the T2 experimental visit was conducted six months post-program completion. Research assistants collecting data were blinded to treatment allocation. However, double blinding was not feasible in this context because the intervention’s nature made it impossible to conceal allocation from the participants.

### Intervention overview

Details on the intervention content have been reported previously.^28,29^ Briefly, the Kiplin intervention is a theory-driven digital APA program including four key components: a) 22 APA sessions (2 face-to-face sessions at the program’s start, followed by 20 remote videoconference sessions), b) three PA collective games accessible through an Android or iOS app, c) chat feature allowing communication with other participants, teammates, and health professionals, and d) an activity monitoring tool enabling participants to track their PA in real-time. The intervention integrates a total of 16 Behavior Change Techniques and primarily aims to promote changes in daily PA behavior.

Participants allocated to the control condition received the traditional 3-month face-to-face supervised APA program at the University Hospital of Clermont-Ferrand, with three sessions a week on non-consecutive days, for a total of 36 sessions. Each session included a warm-up, 50 minutes of endurance and muscle-strengthening exercises, followed by stretching, all conducted under the supervision of an APA coach in a dedicated facility. Further details are available in the study protocol.

### Study outcomes

Full details of the outcome measures, including their reliability and validity, have been reported previously.^28^ At baseline, participants provided demographic data, including date of birth, sex, and highest level of education completed. The primary outcome was the change in daily step count, assessed at high resolution using the Garmin Vivofit 3 (Garmin International, Olathe, Kansas, USA), a wearable activity tracker with validated accuracy under various walking conditions.^30^ Daily step count changes were evaluated from baseline to one-week post-intervention. Additionally, the change in daily steps in the follow-up period from baseline was also evaluated.

The three experimental visits included the following clinical outcomes:

#### Anthropometric measurements and body composition

Height and weight were measured at the same time of day for consistency. BMI was calculated as body mass (kg) divided by height squared (m^2^). Body composition, including fat and lean mass, was assessed using bioelectrical impedance analysis with the multi-frequency segmented body composition analyzer (Tanita MC780, Tanita, Hong Kong, China).

#### PA and sedentary behaviors

Moderate-to-vigorous physical activity (MVPA), light PA (LPA), and sedentary time were measured over seven consecutive days using a tri-axial accelerometer (ActiGraph GT3x; ActiGraph LLC, Pensacola, FL, USA).

#### Physical capacities

Cardiorespiratory fitness was assessed via the 6-minute walking test (6MWT). Muscular strength of the upper limbs was assessed via handgrip strength using a dynamometer (Takei Grip-D, Takei, Japan) with a series of three handgrip test measurements for right and left hands, in the seated position. Muscular strength of lower limbs was assessed through maximum knee extension torque using an isokinetic dynamometer at speeds of 30, 60, and 120°/s. Patients performed two sets of three repetitions for each speed, with the highest value recorded.

#### Quality of life

Quality of life was measured via the EQ-5D-5L questionnaire,^31^ which evaluates five dimensions: mobility, autonomy of the person, current activity, pain/discomfort, anxiety/depression.

#### Program adherence

For both groups, the number of APA sessions attended was assessed. For the Kiplin group, app engagement was collected and comprised participation in the games, app usage frequency, and the number of messages sent.

#### Economic evaluation

The health economic evaluation was conducted through a cost-utility analysis, incorporating (1) the identification and valuation of costs and (2) the measurement of utility using the EQ-5D questionnaire. The analysis was performed from the health insurance perspective, considering only direct medical costs. The time horizon for this evaluation extended from baseline (T0) to the follow-up assessment at T2.

### Statistical analyses

Sample size estimations were based on the primary outcome measure of daily steps measured using the Garmin Vivofit 3. The trial was designed to demonstrate a difference equivalent of an effect size of *d* = 0.6 on our primary outcome, for 80% power and a two-sided type I error at 0.05. In idiographic designs, power is estimated at level 1 (within-person changes) and not at level 2 (number of participants). Recommendations from statistical experts^32^ are for a minimum of 835 observations for adequate power (.80) at level 1. With 2,058 (98 days x 21 participants) observations, we have adequate power to detect within-person changes in the Kiplin condition.^33^

All analyses were performed as a modified intention-to-treat analysis using data from participants with complete data for either the primary or secondary outcomes, at the two first experimental visits. Analyses were performed using R (R Foundation for Statistical Computing, Vienna, Austria). Baseline variables were reported as numbers and percentages for categorical variables and means with standard deviations (SDs) for continuous variables. According to the CONSORT 2010 statement,^34^ group differences in baseline variables were not compared using significance testing.

Changes in daily step count were assessed between baseline and intervention periods, as well as between baseline and follow-up, using linear mixed-effects models. These models accounted for the nested data structure and included fixed effects for group, period (baseline, intervention, or follow-up), and group × period interaction. Random intercepts for participants and random linear slopes for repeated measures at the participant level were included. Adjusted models were further fitted, incorporating fixed effects for group, period, group × period interaction, age, baseline PA levels, BMI, and season—factors previously identified as significant predictors of intervention effects.^29^ Non-wear days, defined as days with fewer than 1,000 steps, were treated as missing data. Model fit was evaluated using the Bayesian Information Criterion (BIC) and -2-log-likelihood (−2LL).^35^ All models were performed using the *lmerTest* package^36^ and contrast analyses were conducted using the *emmeans* package.^37^ Diamond comparison plots were drawn with R package *ufs*.^38^ Contrary to what was outlined in the protocol, group × time interaction analyses for the primary outcome were not conducted due to the potential for nonlinear effects over time. Instead, idiographic analyses were developed to explore the dynamic effects of the intervention across the study period.

In a complementary analysis of the primary outcome, an idiographic approach was employed to analyze daily step counts for each participant separately using Generalized Additive Models (GAMs).^39^ GAMs are an extension of generalized linear mixed models that allow the estimation of smoothy varying trends where the relationship between the covariates and the response is modeled using smooth functions.^40^ GAMs are particularly well-suited to the modeling of time series data with one level of measurement (i.e., repeated measurements nested within one individual), as they can accommodate the inclusion of autocorrelated error terms.^41^ Missing days were imputed using the Kalman Filter method, recommended for univariate time series imputation.^42^ Data were imputed separately for each dataset (i.e., each participant). Nonlinearity was assessed via the effective degrees of freedom (edf) of smoothing terms, with edf ≥3 indicating nonlinearity.^41^ GAMs were computed using the *mgcv* package^43^ and the *visreg* package^44^ was used for model visualization.

Continuous secondary outcomes were analyzed using mixed-effect models (including the group, time, and group x time interaction terms as fixed effects) to assess changes across 9 months (i.e., considering the three measurement points).

## RESULTS

### Participants characteristics

Between June 22, 2021 and October 6, 2022, 57 patients were screened for eligibility, with 50 meeting the inclusion criteria and subsequently randomized to either the Kiplin group (n = 25) or usual care group (n = 25) (Figure 1). Participants who completed at least two assessments (modified-intention-to-treat sample) had a mean age of 47.7 (SD 13.3) years, with 31 women (74%), and a mean BMI of 40.0 ± 7.14 kg/m^2^. A total of 13 participants had a T2D (31%). At baseline, the average daily step count was 6,584 (SD = 3,787) steps per day. No substantial differences in baseline characteristics were observed between groups (Table 1).

**Table 1.** Descriptive statistics of the modified-intention to treat sample.

|  | Kiplin intervention (n = 21) | Usual care (n = 21) | Total (n = 42) |
| --- | --- | --- | --- |
| ***Sociodemographics*** |  |  |  |
| Age, mean (SD) | 47.5 (11.0) | 47.89 (16.13) | 47.7 (13.3) |
| Female, n (%) | 17 (81) | 14 (67) | 31 (74) |
| BMI kg/m<sup>2</sup>, mean (SD) | 39.67 (7.37) | 40.32 (7.02) | 40.0 (7.14) |
| Obese, n (%) | 20 (95) | 19 (90) | 39 (96) |
| T2D, n (%) | 7 (33) | 8 (31) | 13 (31) |
| ***Education*** |  |  |  |
| Less than high school, n (%) | 5 (24) | 8 (38) | 13 (31) |
| High school, n (%) | 6 (29) | 3 (14) | 9 (21) |
| University degree (%) | 10 (48) | 10 (48) | 20 (48) |
| ***Physical activity (daily steps)*** |  |  |  |
| Baseline, mean (SD) | 6,340 (3,269) | 6,827 (4,249) | 6,584 (3787) |
| Missing daily step values, n (%) | 429 (9.5) | 243 (11.1) | 672 (10.3) |

**Figure 1.**
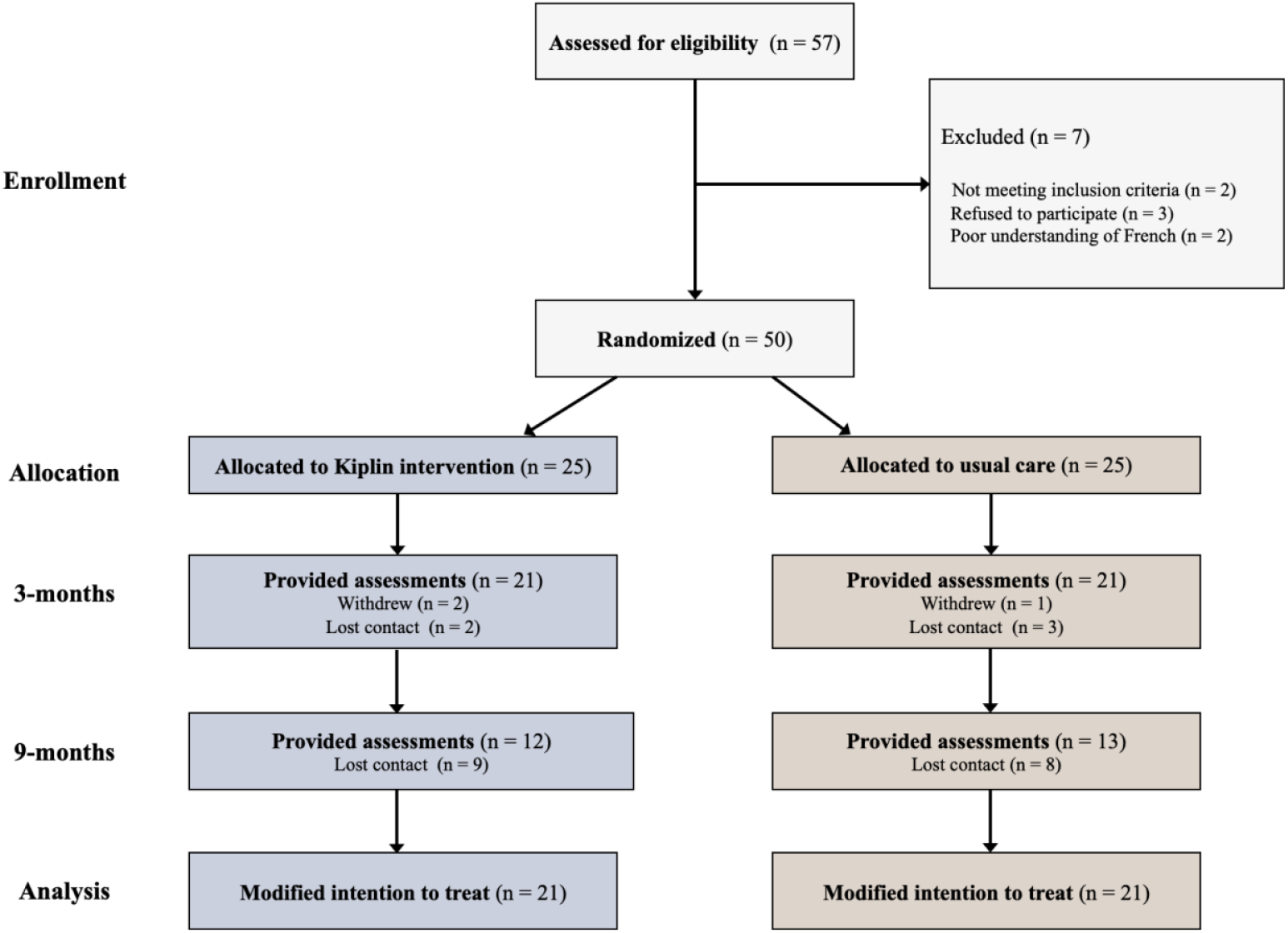
Study flow diagram.

### Compliance and engagement metrics

Among the 50 randomized participants, eight withdrew or were lost before program completion (three in the intervention group and 5 in the control group), and 17 were lost during the follow-up period (Figure 1). Of a possible 274 days, participants logged steps for a median of 158 days (IQR range = 162) throughout the study, with 80% of participants wearing their activity monitor for at least 75% of days. In the Kiplin condition, participants attended an average of 14.68 (SD = 6.14) of 22 possible APA sessions, yielding a completion ratio of 0.67 (SD = 0.28). They engaged in an average of 2.6 (SD = 0.71) of a possible three physical activity games, corresponding to a mean of 36.6 days in-game (SD = 10.02), sent an average of 14.2 messages (SD = 15.93), and logged into the app on 88.12 times on average (SD = 43.63). In contrast, patients receiving usual care attended an average of 30.27 (SD = 5.88) out of 36 APA sessions, with a completion ratio of 0.84 (SD = 0.16).

### Primary outcome

#### Between-group differences

During the 3-month intervention period, participants in the Kiplin condition increased their daily steps by 1092.06 steps per day on average (+10.9%; 95%CI [533; 1651.5], *p*<0.001, *d* = 0.38) compared to the baseline period and by 834 steps per day on average during the follow-up period (+8.3%; 95%CI [114; 1554.7], *p*=0.01, *d* = 0.29) compared to the baseline. In contrast, the daily step count of the usual care condition remained stable and showed no significant changes in daily steps during the intervention period (+0.1%, p=1.00) or follow-up period (+0.9%, *p*=0.10) relative to baseline. Figure 2 depicts the unadjusted daily step counts evolution by phase and condition.

**Figure 2.**
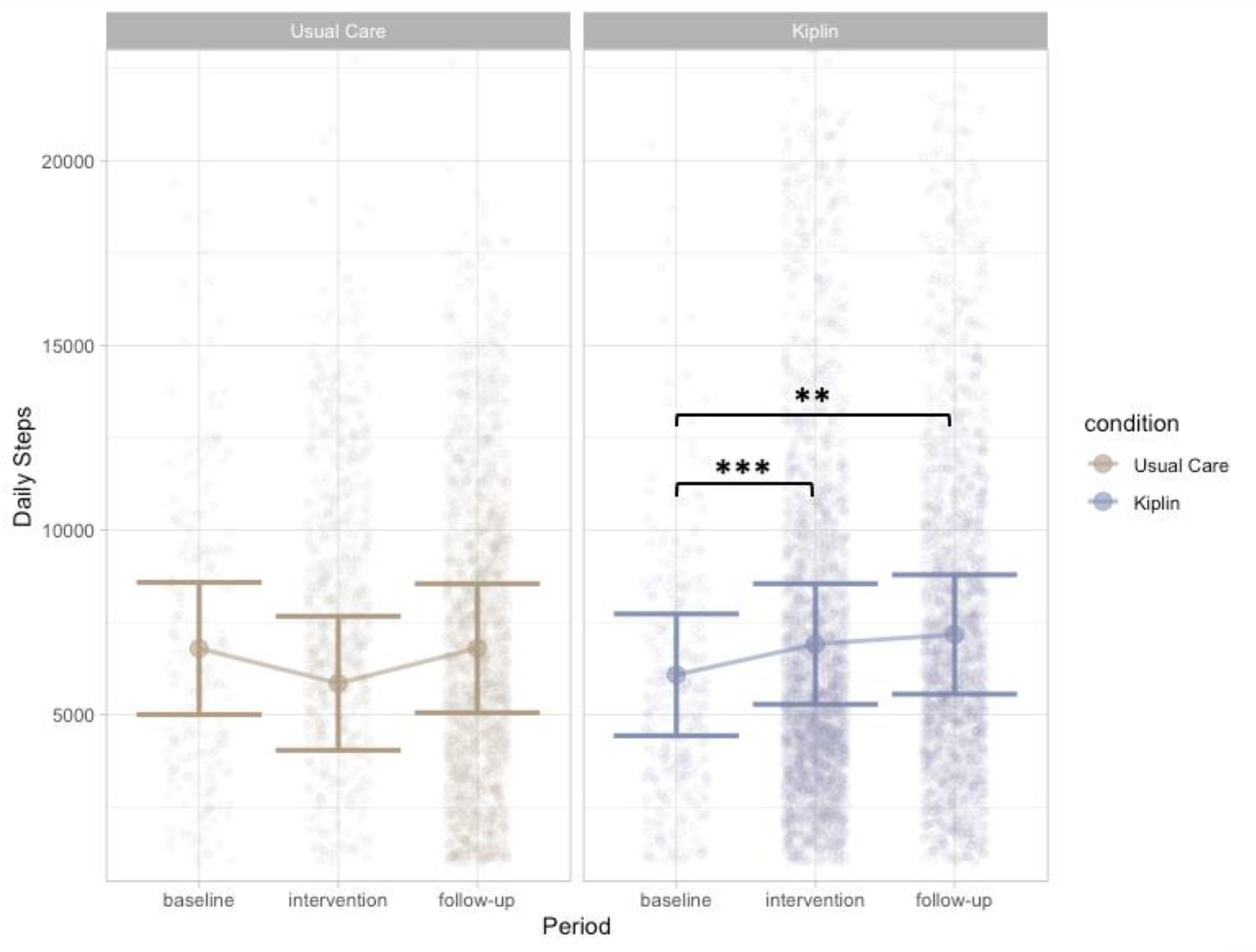
Changes in daily steps throughout the study phases for the Kiplin and usual care conditions. Error bars represent standard error. *** p<0.001; ** p=0.01

Contrast analyses revealed that participants of the Kiplin group had a significantly greater increase in mean daily steps between baseline and the intervention period, compared with the usual care with an estimated difference of 1085 steps (95% CI [493, 1676]; *p*<0.001) relative to baseline. Similarly, during the follow-up period, the daily step count change from baseline of patients in the Kiplin group remained significantly greater of 1775 steps (95% CI [906, 2643]; *p*<0.001).

Results of the adjusted models follow the same trends and are available in Table S1 in supplementary materials.

#### Within-person evolutions

The results of the Generative additive models (GAMs) analyzing the evolution of daily step counts over time for each participant in the Kiplin group are summarized in Table 3. The effective degrees of freedom (edf) for the smoothing terms ranged from 1.0 to 8.10, with 9 participants exhibiting edf > 3, indicative of non-linear patterns. Among the participants, 8 showed significant changes in daily step counts over time (*p* < 0.05), with 7 showing improvements and 1 experiencing a decline. Visualizations of the daily step trajectories for these 8 participants are presented in Figure 3, with the complete set of plots available in the supplementary materials (Figure S1).

**Table 3.** Results of the Generative Additive Models evaluating the evolution of daily steps across time between baseline and one week after the end of the intervention for participants of the Kiplin group.

|  | Estimate | edf | *p* |
| --- | --- | --- | --- |
| Participant #1 | 1.27 | 4.51 | 0.31 |
| Participant #2 | 3.86 | 1.94 | **0.01** |
| Participant #3 | 0.94 | 1.00 | 0.33 |
| Participant #4 | 0.32 | 1.00 | 0.58 |
| Participant #5 | 1.39 | 1.27 | 0.34 |
| Participant #6 | 0.44 | 1.53 | 0.57 |
| Participant #7 | 3.06 | 8.10 | **<.001** |
| Participant #8 | 4.50 | 5.75 | **<.001** |
| Participant #9 | 2.76 | 4.56 | **0.02** |
| Participant #10 | 0.01 | 1.00 | 0.92 |
| Participant #11 | 2.49 | 2.00 | 0.09 |
| Participant #12 | 13.14 | 1.25 | **<.001** |
| Participant #13 | 2.07 | 4.54 | 0.07 |
| Participant #14 | 2.83 | 1.00 | 0.10 |
| Participant #15 | 7.61 | 5.54 | **<.001** |
| Participant #16 | 0.05 | 1.00 | 0.83 |
| Participant #17 | 1.28 | 2.32 | 0.31 |
| Participant #18 | 1.64 | 5.93 | 0.15 |
| Participant #19 | 8.62 | 1.29 | **<.001** |
| Participant #20 | 1.60 | 5.82 | 0.15 |
| Participant #21 | 3.17 | 7.81 | **<.001** |

**Figure 3.**
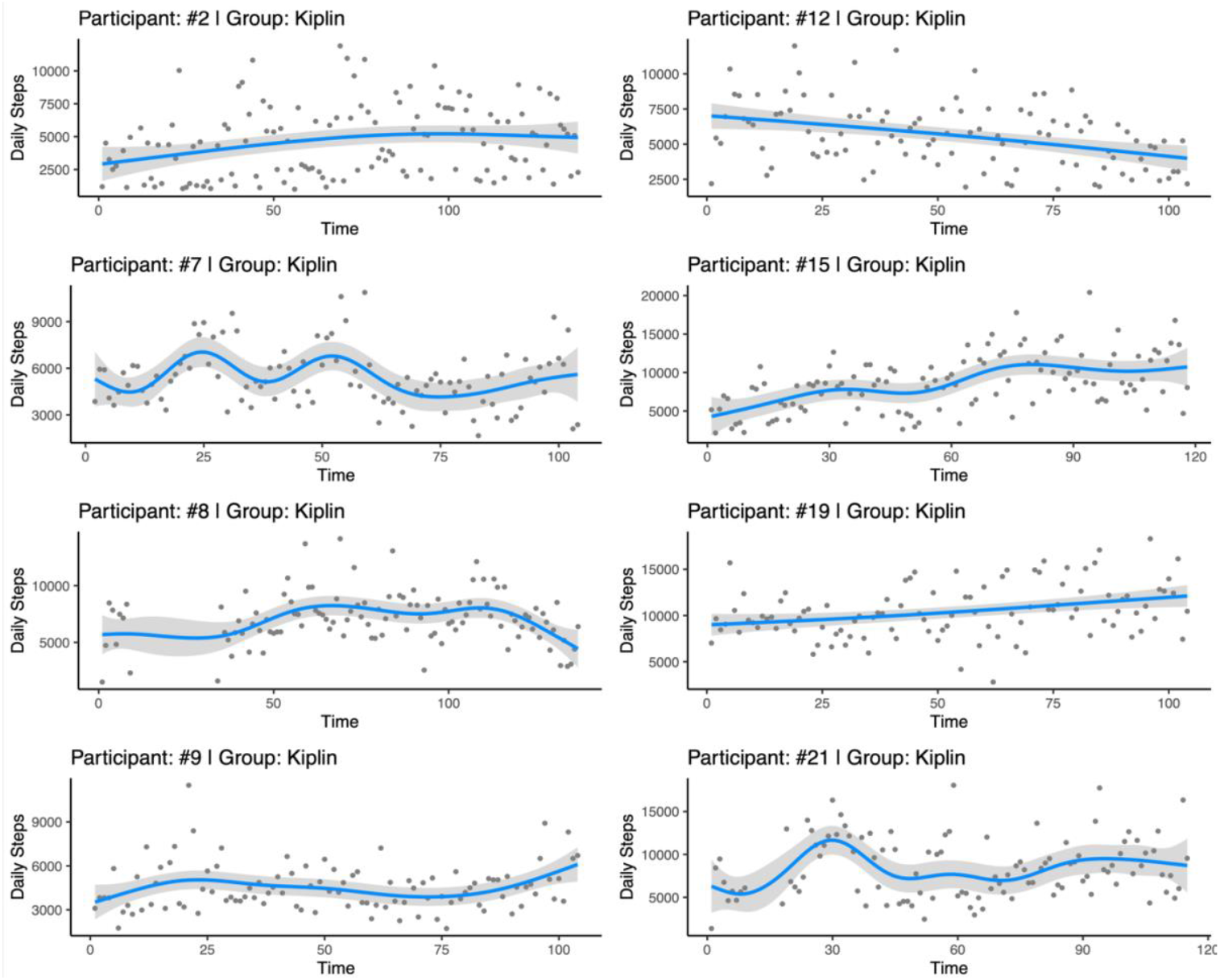
Plots of the Generalized Additive Models for the evolution of daily steps from baseline to one-week post-intervention for the 8 participants of the Kiplin condition with significant changes across time. The plots of other participants are available in Figure S1 of the supplementary materials.

### Secondary outcomes

Table 4 presents the results of the secondary outcome measures. Within-group comparisons showed significant improvements from baseline at 9 months on the lower limb muscle strength (b=6.68, 95%CI [1.40; 11.96], *p=*0.014) and a significant decrease in BMI at 3 months (b=-0.51, 95%CI [-0.98; -0.04], *p=*0.034) for the Kiplin group. In contrast, the control group showed significant improvements in quality of life at 3 months (b=0.08, 95%CI [0.01; 0.15], *p=*0.020) and in 6MWT and lower limb muscle strength at both 3 months (b=38.94, 95%CI [18.96; 58.93], *p<*0.001 and b=9.27, 95%CI [4.15; 14.40,], *p*=0.001 respectively) and 9 months (b=25.41, 95%CI [2.04; 48.78], *p*=0.034, 0.034 and b=7.59, 95%CI [1.80; 13.39,], *p*=0.011 respectively). Results of the mixed-effect models revealed no significant group x time interactions for secondary outcomes, except for a significant interaction in MVPA change from baseline, favoring the Kiplin group (b=5.69, 95%CI [0.61; 10.77,], *p=*0.029). Figure 4 provides a visual comparison of changes between conditions with effect size estimates. Graphs depicting the evolution of secondary outcomes are available in supplementary materials (Figure S2). Although statistical significance was not reached, trends suggest that the Kiplin program had a greater impact on PA and sedentary behaviors.

**Table 4.**
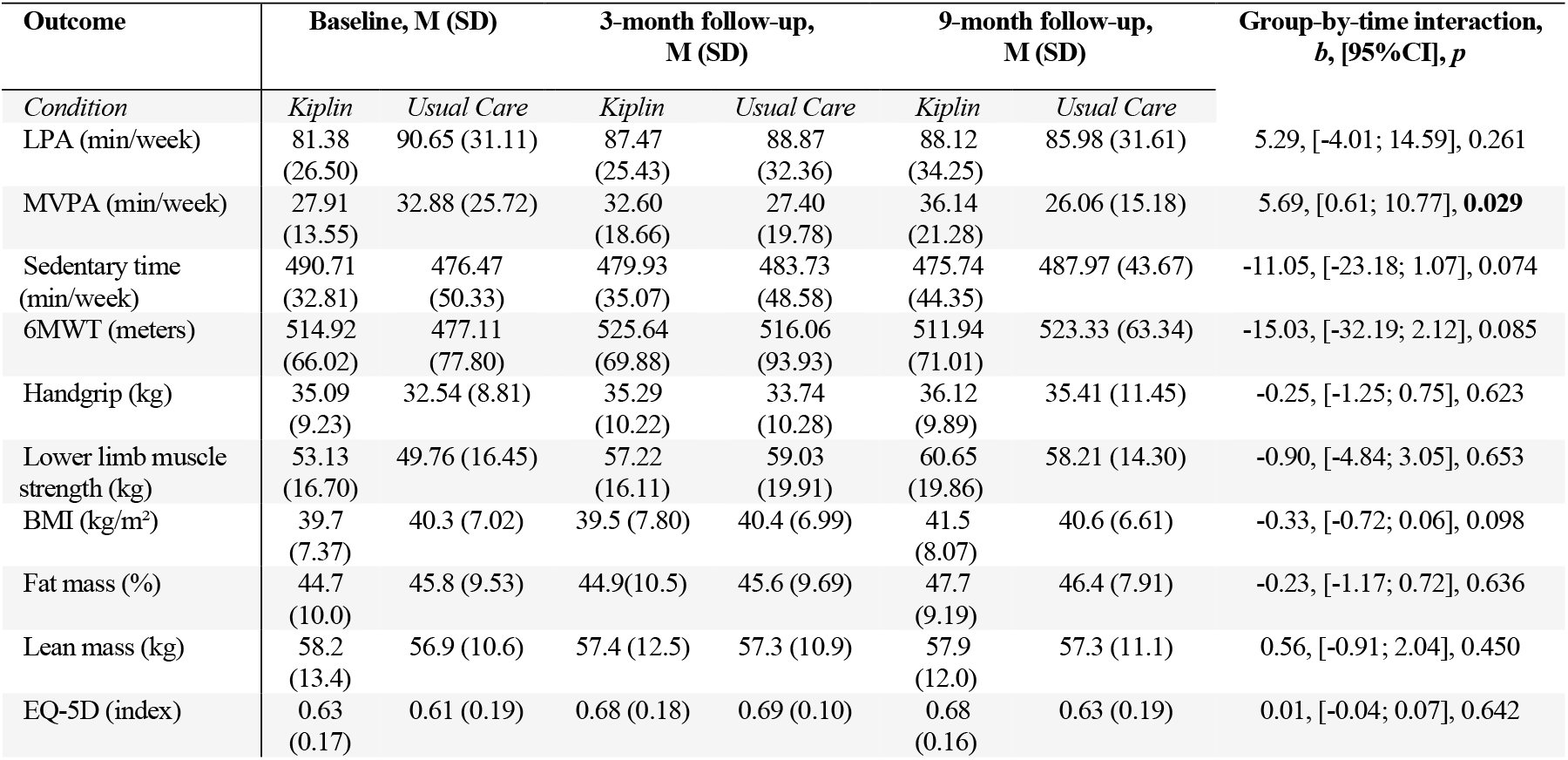
Secondary outcome measures at baseline, 3-, and 9-month follow-up. LPA light physical activity; MVPA moderate to vigorous physical activity; 6MWT 6-minute walking test; BMI body mass index.

**Figure 4.**
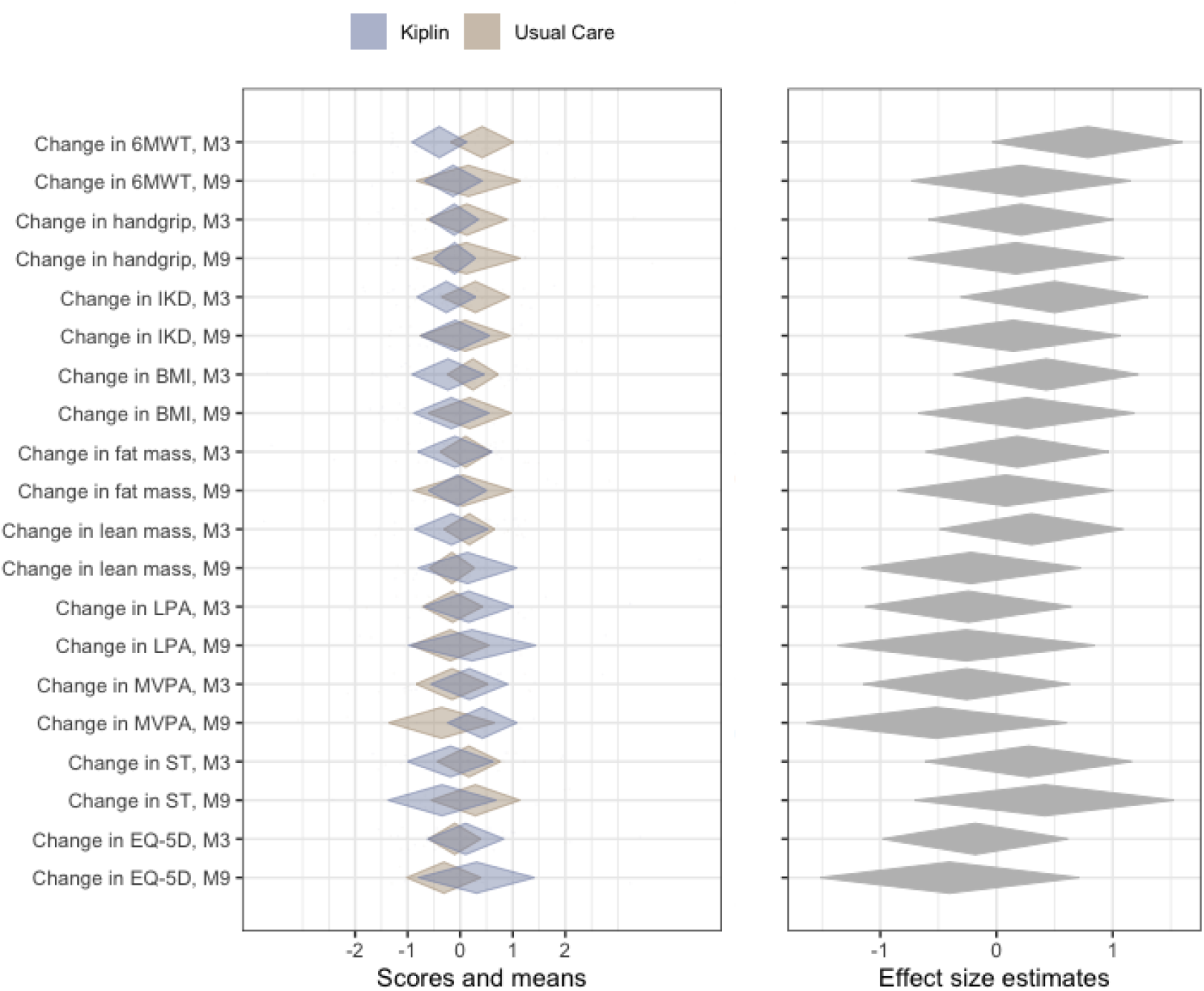
Diamond comparison plot of the univariate standardized changes in the secondary outcomes between the Kiplin and usual care conditions. The middle of the diamonds shows the means and the endpoints of diamonds representing the 99% confidence intervals. 6MWT 6-minute walking test, IKD isokinetic dynamometer, BMI body mass index, LPA light physical activity, MVPA moderate-to-vigorous physical activity, ST sedentary time.

### Economic evaluation

The costs per patient in the usual care group was €491.92 compared to €568.74 in the Kiplin group, resulting in a cost differential (surplus of Kiplin compared to the control group) of €76.82. Quality-adjusted life years (QALYs) did not significantly differ between groups and time points. The QALYs calculated from the EQ-5D questionnaire were, on average, rather good at baseline, with an average of 0.62 at T0 and 0.67 at T1 and T2.

## DISCUSSION

The results of this controlled randomized trial revealed that a group-based gamified digital intervention significantly increased daily PA compared to a traditional supervised face-to-face PA program. Interestingly, this significant difference was observed both during the 3-month program (+1085 daily steps) and during follow-up periods (i.e., 6-month post-intervention, +1775 daily steps). These results remained consistent irrespective of adjustments for potential confounding factors, reinforcing the robustness of the findings. Such findings support previous studies indicating that gamified digital interventions can promote meaningful real-world improvements in daily step counts, lasting several months post-intervention.^14,29,45^

These results are particularly promising for several reasons. Where current traditional face-to-face supervised PA programs often face challenges in driving sustained increases in physical activity,^27^ the Kiplin intervention addresses this gap effectively. Notably, the effect size observed during the intervention was maintained throughout the six-month follow-up, underscoring the sustainability of the PA behavior change in participants with obesity and/or T2D. The benefits of improving daily PA are now well-recognized.^46^ A recent meta-analysis revealed that taking more steps per day was associated with a progressively lower risk of all-cause mortality, regardless of age, health status, or intensity.^47^ This suggests that the observed behavioral changes in the Kiplin group could have significant long-term health implications. These findings also align with recent recommendations to shift the focus of obesity and T2D management from weight loss alone to health-enhancing physical activity, fostering more durable clinical outcomes over time.^48,49^

Nevertheless, in parallel to these results, idiographic analyses revealed substantial variability in individual responses within the Kiplin condition. More especially, GAM models showed a) significant between- and within-person variability during the intervention, with some participants displaying highly non-linear patterns while others showed linear trends, and b) divergent responses to the intervention, with several participants experiencing no significant changes across time. These findings introduce important nuance to our results, indicating that digital interventions may not be suitable for every individual. This underscores the need for effective screening methods to identify patients who are most likely to benefit. Indeed, we can assume that factors such as the stage of behavior change, the acceptability of the technologies,^50^ or some physiological characteristics^51^ could play a critical role in determining whether a digital or in-person program would be more appropriate. This highlights the importance of incorporating the principles of 5P medicine—preventive, personalized, participative, predictive, and evidence-based—into the design and implementation of digital interventions. Furthermore, the observed variability within the intervention emphasizes the need for adaptive designs, such as just-in-time interventions, which can effectively account for the rapidly changing and dynamic nature of behavioral states in physical activity and gamified interventions.^52,53^

In addition, no statistical differences were observed in the secondary outcomes evaluated in this study, except for the MVPA change. While these results should be interpreted with caution, as they stem from secondary analyses potentially without appropriate statistical power, they remain promising from a medico-economic perspective. The Kiplin intervention demonstrated comparable efficacy to usual care with fewer sessions, suggesting potential for cost-efficiency. According to the cost-utility analysis, the cost surplus generated by Kiplin of approximately €76 per patient is not significant for a quality of life that is not impaired. The anticipated gains in QALYS could not be demonstrated, probably due to the limited sample size at T2 and the high baseline quality of life score reported by patients at T0. This suggests that the generic EQ-5D measurement tool, while useful for QALY transposability, may lack the specificity needed to detect quality-of-life changes in this population of patients. To provide a conclusive evaluation of Kiplin’s cost-effectiveness over usual care, future research should conduct more specific medical-economic analyses, incorporating quality-of-life metrics adapted to the patient population. Furthermore, a more comprehensive prospective evaluation of cost differences could include the depreciation of premises during the usual care, which were not factored in this study due to restricted access to detailed institutional financial records. Only direct hospital resource costs were assessed.

From a methodological perspective, the variability observed with the GAMs reflects a broader limitation of traditional RCTs, which typically focus on group-level differences, potentially overlooking meaningful interindividual variability. By relying solely on aggregate measures, RCTs may fail to capture distinct response patterns, limiting insights into underlying mechanisms and moderating factors that influence intervention effectiveness. In this line, idiographic approaches appear as a valuable complement to traditional clinical experimental designs, and this study highlights the importance of combining both between- and within-person approaches for evaluating digital interventions, as well as the advantage of high-resolution behavior measurement. Taking this a step further, high-resolution tracking of clinical outcomes could significantly boost statistical power and decrease measurement inaccuracies. Emerging tools, such as *MediEval* (i.e., an app-based medical device providing valid and reproducible measures of the 6MWT and 30s-STS tests)^54^ or smartphone-derived Seismocardiography (i.e., a method for measuring heart-induced chest vibrations and measuring VO2 max)^55^ could facilitate remote and frequent autonomous assessments of patients’ physical condition. For self-reported measures, the use of Ecological Momentary Assessments (EMA) may be particularly suited.^56^

However, the results should be interpreted in light of several limitations. First, while using wearables allowed for continuous daily step monitoring in real-world conditions, variations in wear time between groups at the day level cannot be excluded, as the Kiplin group may have been more incentivized to wear the device through gamification. Qualitative interviews with the patients suggested that participants in both groups wore the devices consistently, but we lack objective data to confirm this. Future studies using wearables with continuous heart rate tracking could better assess and control for wear time. Second, results from secondary outcomes should be viewed with caution due to potentially insufficient power to detect these effects as the power test was computed for the primary outcome. Lastly, digital interventions consist of a complex interplay of interconnected components, making it challenging to isolate the specific influence of individual elements when the intervention is evaluated as a whole. Consequently, it remains unclear whether the observed behavior change is attributable to the digital format itself or to specific intervention elements. To address this challenge, a more effective approach may involve decomposing the intervention and conducting multi-arm randomized controlled trials to disentangle the effects of gamification elements. In this context, innovative research frameworks like the Multiphase Optimization Strategy (MOST)^57^ or hybrid designs^58^ could offer a promising avenue in future research for systematically optimizing and evaluating digital health interventions.

## Conclusions

This study confirms the potential of digital health interventions to promote sustained changes in PA compared to usual care in participants with obesity and T2D. Although this behavior change has not led to superior clinical outcomes compared to usual care in the present study, its continued persistence beyond the six-month post-intervention period could lead to more pronounced long-term benefits. To validate this hypothesis, larger clinical trials with extended follow-up durations are necessary. This study also had the originality of incorporating both between-group and within-person analyses of daily step counts. The findings indicate that while the digital intervention effectively increased daily steps on average compared to usual care, the benefits were not uniform across all participants. This underscores the importance of patient screening and tailoring program content to individual needs. Future research should further investigate these considerations to optimize digital health intervention design and implementation.

## Data Availability

The anonymized data used in this study and the R code are available on the Open Science Framework (DOI 10.17605/OSF.IO/YPD5S).

https://osf.io/ypd5s/

## Acknowledgments

The authors would like to thank the challenge 3 I-SITE Clermont Auvergne Project 20-25 for their grant, Stéphane Penando and Aliette Wauthier for their involvement in the measurement assignments, Guillaume Harel for his advice on data curation, Dario Baretta and Guillaume Chevance for sharing statistical aspects of time series analyses, and all the included patients for their time.

## Authors contributions

Conceptualization: AM, AC, MB, MD; Methodology: all the authors; Software: AM; Formal analysis: AM; Investigation: AM, MD; Data curation: AM, BP; Writing – original draft: AM; writing – review & editing: all the authors, Visualization: AM; Project administration: AM, MD; Funding acquisition: AM, AC, MB, MD

